# A Cellular atlas of the human fallopian tube reveals the metamorphosis of secretory epithelial cells during the menstrual cycle and menopause

**DOI:** 10.1101/2023.11.22.23298470

**Authors:** M Weigert, Y Li, L Zhu, H Eckart, P Bajwa, R Krishnan, S Ackroyd, RR Lastra, A Bilecz, A Basu, E Lengyel, M Chen

## Abstract

The fallopian tube, connecting the uterus with the ovary, is a dynamic organ that undergoes cyclical changes and is the site of several diseases, including serous cancer. Here, we use single-cell technologies to construct a comprehensive cell map of healthy pre-menopausal fallopian tubes, capturing the impact of the menstrual cycle and menopause on different fallopian tube cells at the molecular level. The comparative analysis between pre- and post-menopausal fallopian tubes reveals substantial shifts in cellular abundance and gene expression patterns, highlighting the physiological changes associated with menopause. Further investigations into menstrual cycle phases illuminate distinct molecular states in secretory epithelial cells caused by hormonal fluctuations. The markers we identified characterizing secretory epithelial cells provide a valuable tool for classifying ovarian cancer subtypes.

**Graphical summary:** Graphical summary of results. During the proliferative phase (estrogen^high^) of the menstrual cycle, SE2 cells (OVGP1^+^) dominate the fallopian tube (FT) epithelium, while SE1 cells (OVGP1^-^) dominate the epithelium during the secretory phase. Though estrogen levels decrease during menopause, SE post-cells (OVGP1^+^, CXCL2^+^) make up most of the FT epithelium.

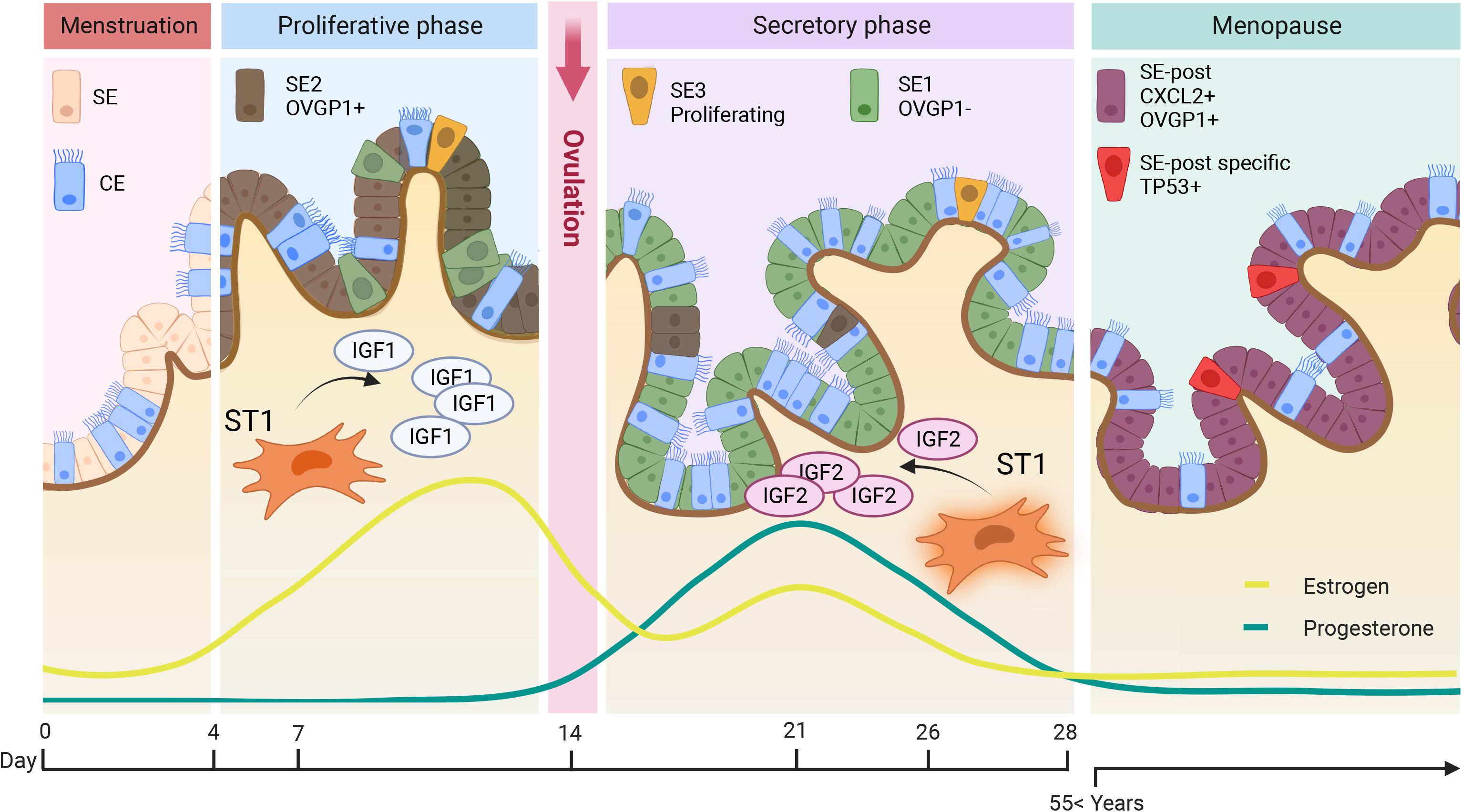

## Introduction

The fallopian tubes (FT) play a vital role in women’s reproductive function. After ovulation, the FT fimbriae pick up the egg from the ovary and muscular contractions and the ciliated epithelial surface cells transport the egg toward the uterus through the ampullary part of the FT, where fertilization takes place. The zygote is then transported towards the narrow intrauterine segment of the FT, from which the blastocyst descends and implants into the uterus (1).

Additionally, the tube serves as an anatomical and immunological barrier against colonization of the abdominal cavity by pathogens. An infection of the tubal lumen (salpingitis) from an ascending infection may lead to occlusion of the fimbriated tubal end, resulting in hydrosalpinx, a high risk of ectopic pregnancy, and tubal factor infertility (2). We know that during menopause, bereft of the protective hormonal environment, the fallopian tube undergoes atrophy (3). However, little is known about cellular changes in the tubes associated with the transition to menopause and if any of these changes predispose women to disease. The fallopian tubes are now recognized as a potential site of origin of high-grade serous cancer (HGSC), since regions of dysplasia with a high proliferation rate, activation of the PI3 kinase pathway, and p53 overexpression are detected in the mucosa of the fallopian tube of women suffering from invasive HGSC (4, 5). Indeed, a mutational analysis comparing p53 mutations in serous tubal intraepithelial carcinomas (STIC) and HGSC showed identical mutations in paired STICs and concurrent HGSC (6).

To better understand the biology of tubal diseases and HGSC, it is crucial to characterize the cellular makeup of the FT throughout the menstrual cycle and menopause. Single-cell technologies have revolutionized our understanding of healthy organs (7) and their associated diseases (8) by allowing the characterization of individual cells revealing their unique gene expression and regulation patterns. Prior single-cell research has led to the recognition of diverse subtypes of FT cells and identified cellular changes that occur with common fallopian tube disease (9–13). However, this research has not systematically compared cellular diversity, gene expression, and regulation of these cells in different phases of the menstrual cycle or by menopausal status. By leveraging single-cell gene expression and chromatin accessibility assays, we deciphered the molecular transformations occurring in specific cell types during the menstrual cycle and menopause. We generated a high-quality fallopian tube cell atlas that allowed us to examine the physiological changes and cellular functions that occur in the human FT during aging. This atlas provides insights that will substantially improve our understanding of diseases affecting the fallopian tubes.

## Results

### Cohort description and fallopian tube collection

The cohort for this study included 17 fallopian tubes (FT) from healthy female donors (designated as D1-7, and D9-D18; **Table S1**) who underwent surgery to treat vaginal prolapse and did not show any major macroscopic or histologic abnormalities in their FT (**Fig. 1A**). We analyzed the different anatomic regions, including the isthmus, ampulla, and fimbriae (**Fig. 1A**), using single-cell RNA sequencing (scRNA-seq) and single-cell chromatin accessibility analysis (scATAC-seq).

**Figure 1:**
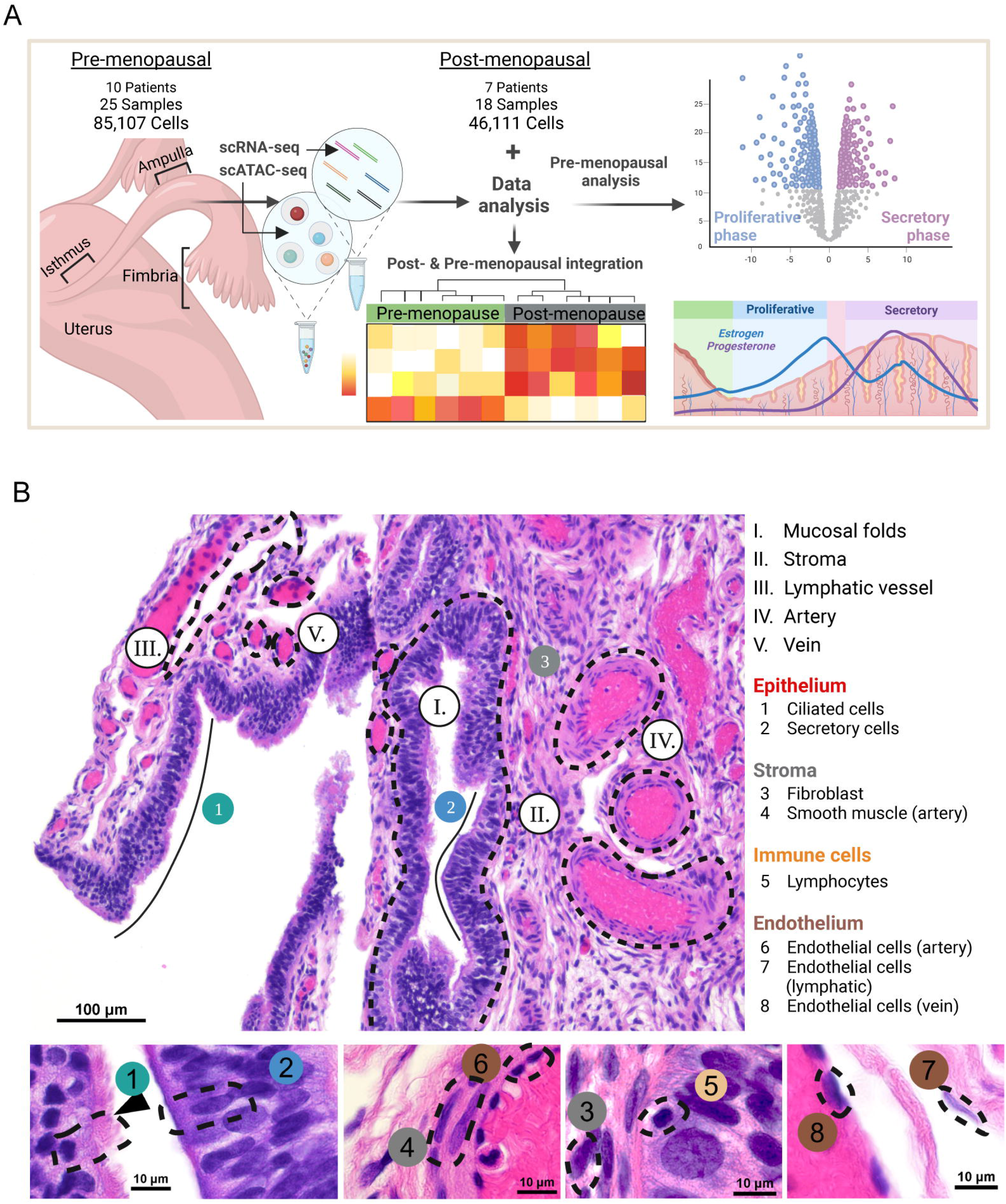
Study design and fallopian tube histology. A) Pre-menopausal fallopian tube (FT) fimbria (F; n = 10), ampulla (A; n = 10) and isthmus (I; n = 5) were characterized using scATAC-and scRNA-Seq. The cellular and molecular changes driven by the menstrual cycle and by menopause in the normal human fallopian tube were integrated. B) Hematoxylin and eosin staining of a normal pre-menopausal FT fimbria. Dashed lines and roman numerals indicate principle cell compartments defined by H&E in the FT. Black lines indicate stretches of ciliated (1) or secretory (2) epithelial cells, and (3) stromal cells. Different cell types identified in the FT by H&E at higher magnification. Specific cell types are marked with circles or by an arrowhead and annotated by numbers.

The results of the seven post-menopausal samples (D1-D7) were previously reported (11). For this study we collected the same FT anatomic regions from an additional ten pre-menopausal donors (D9-D18). We expeditiously transported fresh tissues from the operating room to the laboratory for cell dissociation, followed by scRNA-seq and scATAC-seq. After removing doublets and cells with high mitochondrial content, 85,107 cells from pre-menopausal donors (ampulla and fimbriae only) were integrated with the post-menopausal donors (46,111 cells) before further analysis.

### Cellular composition of the pre-menopausal human fallopian tube

After scRNA-seq and bioinformatics analysis (**Fig. 1A**), we identified 19 clusters across all pre-menopausal samples using unsupervised clustering (**Fig. 2A**, **Table S2**). Based on the expression of canonical marker genes (**Table S3**), the 19 FT clusters were classified into 12 major cell types (**Fig. S1A-B**): ciliated epithelial (CE), secretory epithelial (SE) 1-3, smooth muscle (SM), pericyte/vascular (P/V) 1-2, endothelial (EN) 1-2, lymphatic endothelial (LE), stromal (ST) 1-3, mast (MA), T and natural killer (NK) cells, macrophages (MP), monocytes (MN), B and plasma (B/P) cells (BP) 1-2.

**Figure 2:**
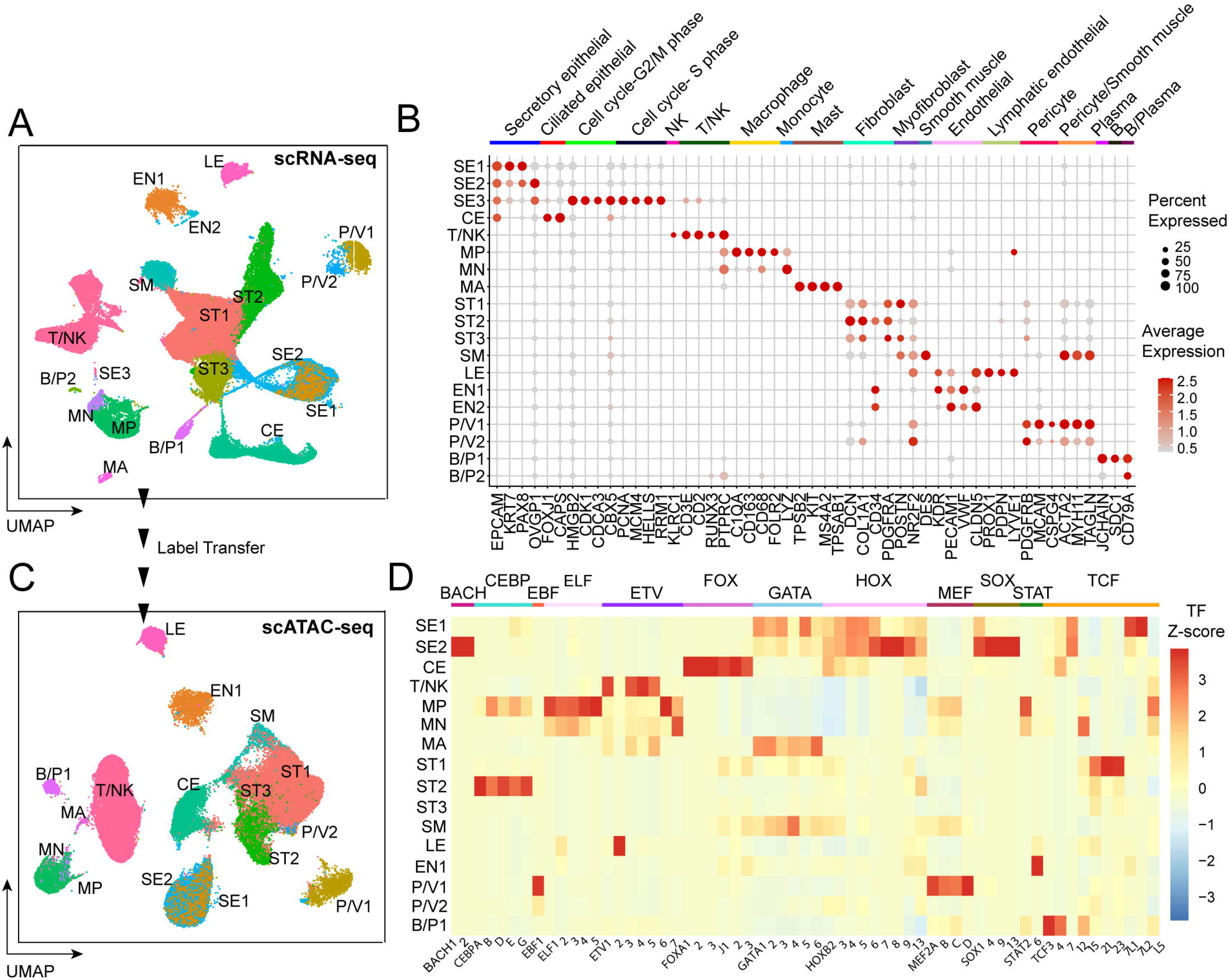
Cellular composition of the normal pre-menopausal fallopian tube. A) Canonical cell types and subtypes found in the ampulla (n = 10) and fimbria (n = 10) of the normal pre-menopausal fallopian tube. The UMAP plot visualizes the 19 cell clusters identified using scRNA-seq. Abbreviations: SE1-3 = 3 subtypes of secretory epithelial cells; CE = ciliated epithelial cells; T/NK = T and natural killer cells; MP = macrophages; MN = monocytes; MA = mast cells; ST1-3 = 3 subtypes of stromal cells; SM = smooth muscle cells; LE = lymphatic endothelial cells; EN1-2 = 2 subtypes of endothelial cells; P/V1-2 = 2 subtypes of pericytes and vascular smooth muscle cells; B/P1-2 = 2 subtypes of B cells and plasma B cells. B) Dot plot showing normalized gene expression levels of canonical marker genes associated with common cell types identified in the dataset (scRNA-seq). C) The UMAP plot visualizing the 16 cell clusters identified in pre-menopausal ampulla (n = 5) and fimbria (n = 5) using scATAC-seq. Cell type labels were transferred from scRNA-seq data and match abbreviations described in A). Cell types SE3, EN2 and B/P2 were not identified in this analysis. D) Heatmap showing transcription factor activity enrichment by cell type based on scATAC-seq data.

Several cell types contained distinct subtypes. Within the SE cell type, we found three subtypes. SE1 and SE2/3 are distinguishable by different levels of *OVGP1* expression and SE3 cells are characterized by the expression of cycle genes (**Fig. 2B**). Though rare, we observed SE3 in all pre-menopausal samples (58 cells detected, **Table 4**). We separated ST cells into three subtypes: ST1 cells express the myofibroblast markers *POSTN* and *NR2F2*, ST2 cells feature high expression of *CD34*, while ST3 cells express canonical fibroblast and myofibroblast markers (*COL1A1*, *POSTN*). We also separated EN cells into two clusters based on the expression patterns of *KDR* and *CLDN5*. The two subtypes of P/V differ in the expression levels of canonical pericyte markers *MACM*, *ACTA2*, *MYH11*, and *TAGLN*, which are considerably lower in P/V2 than in P/V1. However, P/V2 show strong *NR2F2* expression. Lastly, B/P cells comprise two subtypes: B/P1 expresses B and plasma cell markers, while B/P2 expresses only *CD79A* (**Fig. 2B**).

We performed scATAC-seq (10X Genomics) on a subset of pre-menopausal samples and matched them to their corresponding scRNA-seq data (**Table S1**). The cells from ampulla and fimbriae from 10 donors were processed separately and integrated *in silico*. For each sample, we annotated its cluster identity by transferring labels from its scRNA-seq. Specifically, using the scRNA-seq cluster information as a reference, each cell in scATAC-seq was annotated by searching for the best-matched cluster from the same sample. We identified 68,152 cells from the pre-menopausal FT (**Fig. 2C** and **S1C**) from scATAC-seq with matched scRNA-seq compartments (**Table S5)**. The majority of scRNA-seq and scATAC-seq clusters matched well, confirming the high quality of both datasets. However, SE3, EN2 and B/P2 cell subtypes were not detected in scATAC-seq.

To characterize the regulatory landscape in pre-menopausal FT, we assessed the activities of 870 transcription factors (TF) listed in the *cisBP* database (14) across all major cell types (**Fig. S1D**, **Table S6**). Our analysis revealed notable cell-type–specific TF activities, such as strong TCF family activity in B/P1, STAT activity in EN1, FOX family members in CE, ETV family in T/NK, and GATA activity in MA. Our TF enrichment analyses identified differences in TF activities in cell subtypes, further corroborating our decision of subtyping these major cell types. For instance, SE1 shows higher accessibility to GATA, TCF, HOX TF, while HOX, SOX, and the transcriptional repressor BACH accessibility characterized SE2. ST1 and ST2 show differences in the activities of both the TCF and CEBP families; and the activities of EBF1 and the MEF family vary between P/V1 and P/V2 (**Fig. 2D**).

### Human pre-menopausal and post-menopausal fallopian tubes differ in cellular composition and gene expression

We compared the cellular composition of our pre-menopausal FT samples with the post-menopausal FT samples analyzed previously (11). The results indicate that the cellular composition and gene expression of the FT undergoes dramatic changes during menopause (**Fig. 3A**). Among all cell types, the abundance of CE, SE, and T/NK cells decreased significantly in post-menopausal women, while the abundance of EN1, EN2, P/V1, and P/V2 cells increased. There was a trend of more stromal cells (ST2, ST3) in postmenopausal women, but the observed variability was high between samples. While we observed dramatic changes in cell composition, canonical cell marker expression did not change significantly during menopause (**Fig. S2A**).

**Figure 3:**
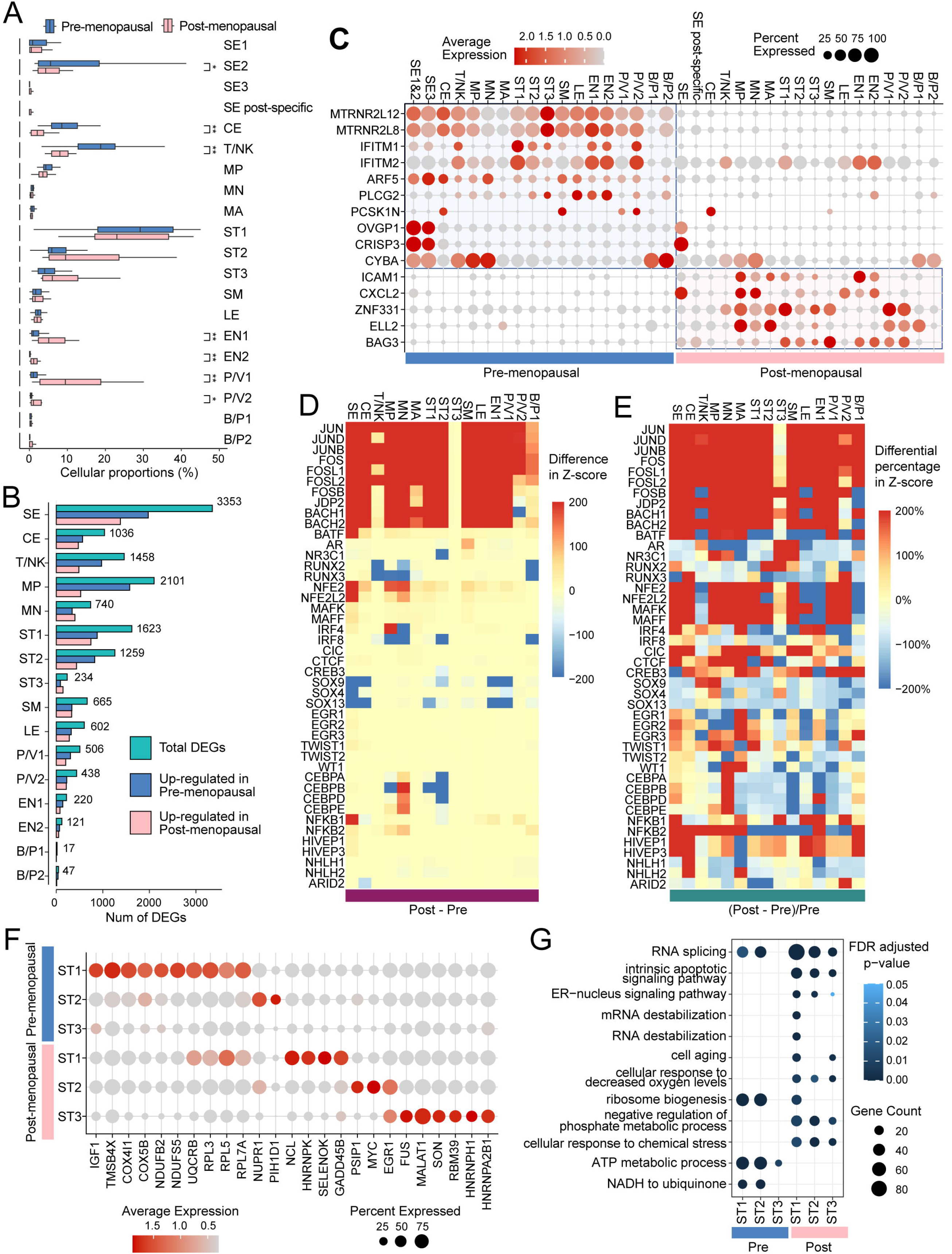
Menopause changes the cellular composition and gene expression profiles of cell types found in the normal human fallopian tube. A) Differences in cellular composition between pre- (n = 11) and post-menopausal (n = 7) women in percentages for combined anatomical sites (ampulla and fimbria). Two-sided t-test. * p-value < 0.05, ** p-value < 0.01. B) Number of differentially expressed genes in pre- and post-menopausal women by cell type using scRNA-seq. Number of DEGs is for ampulla and fimbria combined. C) Dot plot showing normalized gene expression levels of selected, differentially expressed genes specific to menopausal status by individual cell types (scRNA-seq). D) Heatmap showing selected differential transcription factor (TF) activities by cell types using scATAC-seq. The difference in z-score was calculated by subtracting the average pre-menopausal TF activity from post-menopausal TF activity. E) Heatmap showing altered activities of selected transcription factors (TF) by cell types using scATAC-seq data. The differential percentage in Z-score is expressed as percentages relative to pre-menopausal TF activities. F) Dot plot showing normalized gene expression levels of selected genes in pre- and postmenopausal ST subtypes (scRNA-seq). G) Dot plot showing the enrichment of selected gene ontology (GO) terms for each ST subtype in pre- and post-menopausal women based on scRNA-seq data (FDR-corrected p-value < 0.05).

To understand the changes in gene expression profiles between pre- and post-menopausal fallopian tubes, we conducted cell-type–specific differential gene expression analysis using pseudo-bulk donor-level data. Among all cell types, SE cells exhibited most differentially expressed genes (DEGs) (**Fig. 3B**, **Table S7**), followed by MP, ST1, T/NK, ST2, and CE. Several genes were specific to menopausal status. *PCSK1N*, *OVGP1*, *CRISP3*, and *CYBA* (involved in phagocytosis), exhibited significantly higher expression levels in pre-menopausal compared to post-menopausal donors (**Fig. 3C**). Pre-menopausal donors exhibited expression in *IFITM1*/2 and *PLCG2*, which are critical for viral defense. Post-menopausal donors expressed senescence and aging markers such as *ICAM1*, *CXCL2*, *ZNF31*, *ELL2*, and *BAG3* (**Fig. 3C** and **S2B**). Next, we investigated changes in the frequency of interactions among different cell types in the pre- and post-menopausal ampulla and fimbria as detected by *CellPhoneDB*. We observed a reduction in stromal and immune cell interactions after menopause (**Fig. S2C**).

Comparing cell-type–specific activity scores of TFs in post-menopausal FT to pre-menopausal levels, the JUN and FOS families of TFs showed significantly higher activities after menopause in all cell types except ST3 (**Fig. 3D**). Similarly, the BACH1 and BACH2 TFs displayed better chromatin accessibility in post-menopausal samples for all cell types except ST3, P/V1, and P/V2 (**Fig. 3D**, **Table S8**). To avoid overlooking TFs with high relative changes but low absolute changes, we assessed the percentage of change in activities relative to their pre-menopausal levels (**Fig. 3E**). In post-menopausal donors, the androgen receptor (AR) and the glucocorticoid receptor (NR3C1) exhibited an increase in ST3 and SM; the estrogen receptor (ESR) family activities increased in SE1 and MA; TWIST1, an EMT marker increased in ST1 and MP (**Fig. 3E**).

During menopause, stromal cells in the fallopian tubes undergo significant changes that leads to thinning of the FT wall and a reduction in fold complexity (15). Our analysis revealed a molecular upheaval associated with these physiological changes (**Fig. 3F**). Prior to menopause, ST1 cells showed high expression levels of genes such as *IGF1*, *TMSB4X*, *COX5B*, *COX411*, *NDUFB2*, *NDUFS5*, and ribosomal protein coding genes, which drastically decreased in post-menopausal samples. Similarly, *NUPR1* and *PIH1D1* expression were high in pre-menopausal ST2 cells but decreased in menopause. Conversely, the expression of *NCL*, *HNRNPK*, *SELENOK*, and *GADD45B* surged in ST1 after menopause. Likewise, *PSIP1* and *MYC* surged in ST2, and *FUS*, *MALAT1*, *SON*, *RBM39*, *HNRNPH1*, and *HNRN2B1* surged in ST3. Additionally, the expression of *EGR1* increased drastically in both ST2 and ST3 after menopause (**Fig. 3F**). The GO enrichment analysis (**Fig. 3G**, **Table S9**) of the top expressed genes in each ST group revealed that pre-menopausal STs are enriched in genes required for metabolic activity (e.g., ribosome biogenesis, ATP metabolism). In contrast, post-menopausal STs are enriched in genes involved RNA destabilization, cell aging, and apoptosis (**Table S10**).

### Secretory epithelial cells change more significantly than other cell types between pre- and post-menopausal donor samples

We have previously studied the cellular makeup of fallopian tubes from post-menopausal donors. Our analysis of the differences between pre- and post-menopausal samples, uncovered a distinct cluster of cells, unique to the post-menopausal cohort. We named this cluster "SE post-specific" to differentiate it from the remaining post-menopausal SE. Similarly, SE3-pre was found exclusively in pre-menopausal samples and was not detected in any post-menopausal samples (**Fig. 4A** and **S3A**).

**Figure 4:**
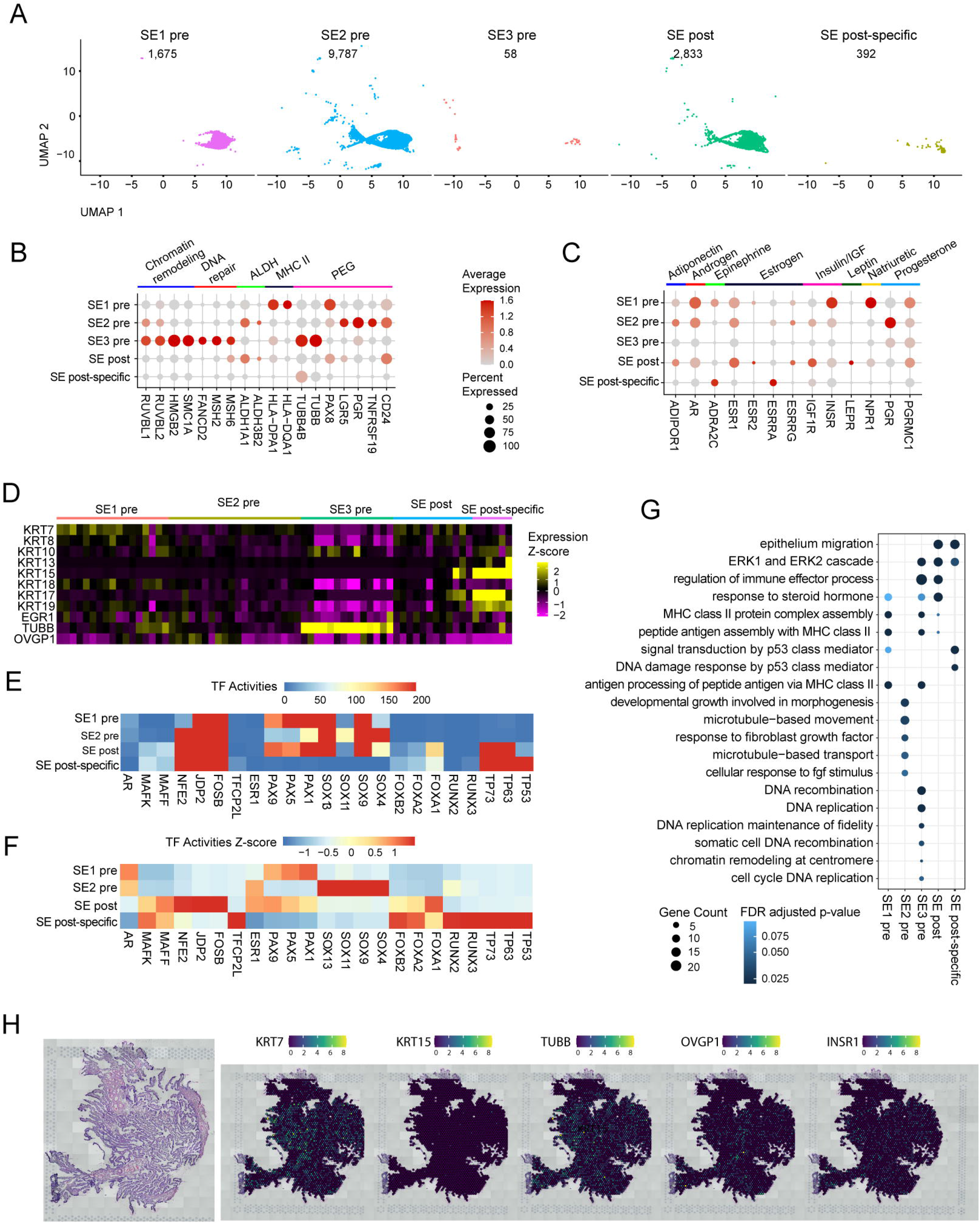
Menopause induces significant changes in gene regulation and -expression in epithelial cells. A) pre- and post-menopausal SE cells identified using UMAP profiling. The pre-menopausal fallopian tube (n = 10) is characterized by three SE subtypes (SE1 pre, SE2 pre, and SE3 pre), while the post-menopausal fallopian tube (n = 7) is characterized by two SE subtypes (SE post and SE post-specific). The number of cells identified in each subtype is shown in the panel (scRNA-seq). B) Dot plot showing normalized gene expression levels of selected markers associated with proliferation, stemness, and PEG-cells in pre- and postmenopausal SE subtypes based on scRNA-seq data. C) Dot plot showing normalized gene expression levels of hormone receptors in pre- and postmenopausal SE subtypes based on scRNA-seq data. D) Heatmap showing the average expression of target genes in SE pre- and postmenopausal subtypes at the individual sample level based on scRNA-seq data (ampulla and fimbria). E) Heatmap showing selected transcription factor activities in SE subtypes based on scATAC-seq data. F) Heatmap showing relative transcription factor activities in SE subtypes using scATAC-seq data. The Z-score is obtained by centering and re-scaling across SE subtypes for each TF. G) Dot-plot showing the enrichment of selected gene ontology (GO) terms for each SE subtype (scRNA-seq). H) Spatial transcriptomics profiling of normal pre-menopausal fallopian tube fimbria using 10x genomics Visium. Pre-menopausal fallopian tube fimbria were analyzed for spatial expression of KRT7, KRT15, TUBB, OVGP1 and INSR1.

In pre-menopausal subtypes (**Fig. 4B** and **S3B**), SE1 pre is characterized by increased expression of MHC class II proteins (*HLA-DPA1, HLA-DQA1)* and *PAX8, a* known lineage marker for high grade serous ovarian cancer used as a clinical biomarker (16). In contrast, SE2 pre is defined by high expression of PEG cell markers (*LGR5*, *PGR*, *TNFRSF19*, *CD24)*, which are a non-ciliated secretory cell type secreting mucus required for decapacitation of spermatozoa before fertilization (17). Lastly, SE3-pre is distinguished by chromatin remodeling markers (*RUVBL1*, *HMGB2*, *SMC1A*), DNA repair markers (*FANCD2*, *MSH2*, and *MSH6*), and *TUBB*. In contrast, post-menopausal SE cell clusters exhibited decreased expression levels of PEG, MHC II, DNA repair and chromatin remodeling markers when compared to pre-menopausal SE cells (**Fig. 4B** and **S3B**). This further indicates that SE cells undergo significant changes during menopause.

Pre- and post-menopausal SE cells also differ in their hormone receptor expression (**Fig. 4C** and **S3C**). In pre-menopausal samples, SE1-pre cells strongly express *AR*, *INSR,* and *NPR1*, while SE2-pre is characterized by its strong expression of progesterone receptor (*PGR*), and SE3-pre displays moderate expression in PGR and PGR membrane component 1 (*PGRMC1*). In post-menopausal SE cells, the overall expression levels of hormone receptors are decreased but a small proportion of cells highly expressed *ESR1*, *ESR2*, *IGFIR* and *LEPR*.

Keratins (KRT) are intermediate filament proteins that provide mechanical support and organization to epithelial tissues (18). We found that KRTs 13, 15 and 17 are associated with menopause, and are only expressed in SE post-specific cells (**Fig. 4D**).

The TF enrichment analysis indicated that there were major differences in TF activity among the different SE groups (**Fig. 4E-F**). The SOX family, involved in tissue repair and regeneration (19), showed activities in all pre-menopausal SE groups, but with highest activity in SE2-pre. PAX-1/5/9 were strongly expressed in SE1-pre and SE-post, consistent with their role in regulating SE gene expression (20). In contrast, TP-53/63/73 were active only in post-menopausal samples, with SE post-specific, having strong expression of TP53 (**Fig. 4E-F** and **S3D-E**). Additionally, SE post-specific cells demonstrated the highest activities in TFCP2L, FOX-A1/A2/B2 and RUN-X2/X3. AR activities decreased while proto-oncogenes from the MAF TF family show strong chromatin accessibility after menopause (**Fig. 4E-F**).

Gene ontology (GO) enrichment analysis of the top expressed genes in each SE group suggests a specific function for each cell cluster (**Fig. 4G**, **Table S9**). SE1-pre is hormone responsive and interacts with immune cells. SE2-pre, expresses genes activated by fibroblast growth factor and microtubule function. SE3-pre expresses many genes involved in DNA replication and immune effector processes. Post-menopausal SE cells are enriched in genes necessary for migration but also hormone response (21), while SE post-specific exhibits enrichment in genes required for the p53-mediated response to cellular stress.

To validate the scRNA-seq findings, we performed full spatial transcriptomics (Visium) on independent pre-menopausal samples. Using *KRT7* as a marker for the SE pre-menopausal cells, we found that *TUBB*, *OVGP1*, and *INSR* co-localized with *KRT7*-expressing areas, providing additional evidence for their role in the pre-menopausal functions of SE cells. As expected, we did not detect expression of *KRT15* in any of the analyzed cells (**Fig. 4H**).

### During the menstrual cycle, secretory epithelial cells change more than other cell types

To examine gene expression changes during the proliferative and secretory phases of the menstrual cycle (**Fig. S4A**) (22), we isolated cells from donors with normal menstrual cycles (25 – 32 days). Our initial observation revealed that SE2-pre cells were predominantly identified in samples from the proliferative phase and marked by high *OVGP1* expression, whereas SE1-pre cells were mostly found in the secretory phase and lack *OVGP1* expression (**Fig. 5A** and **S4B**). To validate the menstrual-cycle–specific expression of *OVGP1* in SE cells, we performed IHC on patient-matched samples. Strong OVGP1 protein staining emerged in the FT epithelium during the proliferative phase but decreased during the secretory phase (**Fig. 5B**). We also validated the post-menopausal expression of OVGP1 and confirmed that SE1-pre cells are regulated by progesterone (**Fig. S4B-C**) while SE2-pre cells and *OVGP1* expression are estrogen regulated (**Fig. 5C-D** and **S4B**), consistent with scRNA-seq and ATAC-seq results of SE1-and SE2-pre (**Fig. 3D-F**). This suggests that they represent distinct cellular states rather than separate cell types, with each state being associated with a specific phase of the menstrual cycle.

**Figure 5:**
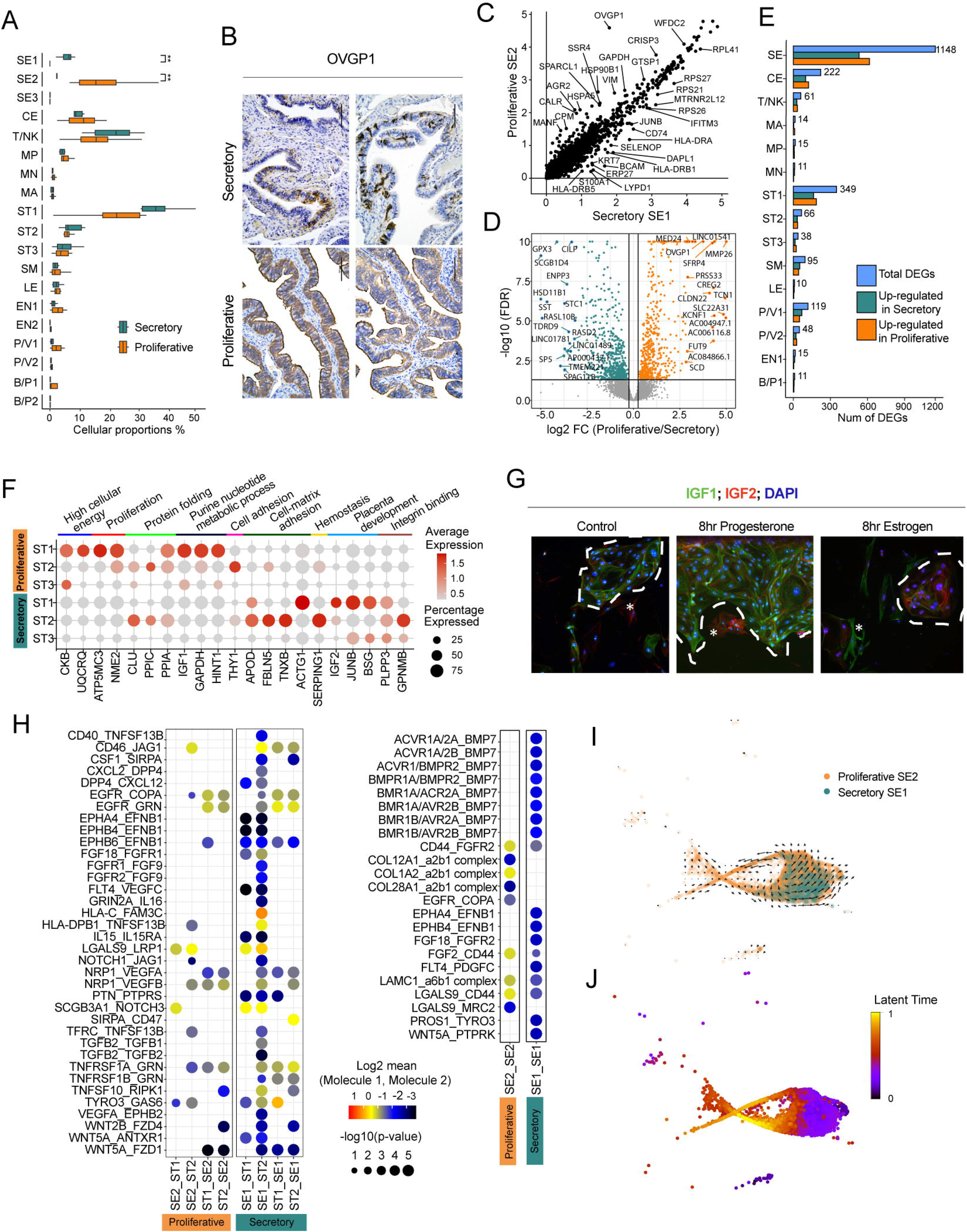
Epithelial cells undergo significant cellular and molecular changes during the proliferative and secretory phases of the menstrual cycle. A) Differences in cellular composition between the proliferative-(n = 4) and secretory phases (n = 3) of the menstrual cycle based on scRNA-seq data (two-sided t-test. * p-value < 0.05, ** p-value < 0.01). B) OVGP1 immunohistochemistry in the fallopian tube of patients in proliferative phase (n = 2) and in secretory phase (n = 2). C) Scatter plot comparing gene expression levels of pre-menopausal SE1 (secretory phase) and SE2 cells (proliferative phase) using pseudo-bulk RNA analysis. D) Volcano plot derived from pseudo bulk analysis. The volcano plot is showing the differential gene expression analysis of genes expressed in SE cells (SE 1/2/3 pre) based on the proliferative and secretory phase. E) Number of differentially expressed genes between the proliferative-and secretory phases of the menstrual cycle by cell type based on scRNA-seq data. F) Dot plot showing normalized scRNA-seq derived gene expression levels of selected genes that differ in the proliferative and secretory phase in ST subtypes. G) Immunofluorescence of IGF1 and IGF2 in primary human stromal and epithelial fallopian cell co-culture. The co-culture was stimulated for 8 hours with either 500nM of estrogen (E4) or 500nM of progesterone (P4). H) Ligand-receptor interactions, detected by CellphoneDB, between SE and ST subtypes (left) and between SE cells (right) separated by the menstrual phase using scRNA-seq data. I) RNA velocity analysis of SE1 and SE2 cells. Arrows indicate the location of the estimated future cell state. Long vectors mark rapid transition events (i.e., large changes in gene expression), while short arrows indicate homeostasis. J) Median latent time for SE cells during the proliferative-and secretory phase of the menstrual cycle.

SE cells displayed the greatest number of DEGs in both menstrual phases (**Fig. 5E**), followed by ST1 cells. Through this analysis, we discovered novel markers for SE1-pre and SE2-pre cells (**Fig. 5D**, **Table 11**), in addition to the markers identified using cell-level analysis (**Fig. 5C**). During the proliferative phase, DEGs in SE2-pre are predominantly associated with COPI-coated vesicle functions transporting cargo between the Golgi and the endoplasmatic reticulum (23). Conversely, in the secretory phase, SE1-pre shows DEGs enriched in clathrin-coated vesicle functions (**Fig. S4D**). Similarly, in ST1 cells, elevated energy consumption and protein folding are linked to the proliferative phase, whereas integrin binding and cell-matrix adhesion are associated with the secretory phase (**Fig. 5F**). There was inverse expression of *IGF1* and *IGF2* in ST1 cells during the menstrual phases (**Fig. S4E**).

A co-culture of primary human stromal and epithelial cells from patients operated for benign conditions was stimulated with progesterone which induced *IGF2* protein expression, while estrogen treatment (E2) induced *IGF1* (**Fig. 5G**).

We further investigated the alterations in receptor:ligand interactions during the menstrual cycle phases by utilizing the *CellPhoneDB* database. In pre-menopausal SE cells, the proliferative phase was marked by increased communication between collagen molecules and the integrin α2β1 complex, while we observed a significant increase in *BMP7* and *EFNB1* interactions during the secretory phase (**Fig. 5H**). Overall, the proliferative phase had fewer receptor:ligand interactions than the secretory phase (**Fig. 5H** and **S4G**).

RNA velocity analysis utilizes both un-spliced (nascent) and spliced (mature) mRNA molecules to predict potential directionality and the time it takes to transition between different cell states (24, 25). We overlaid the velocity vectors in UMAP space to reveal the direction of cellular transitions between SE1-and SE2-pre during the menstrual cycle. The RNA velocity vectors in pre-menopausal SE cells formed a circle, suggesting it being part of a cyclic biological process (**Fig. 5I**). This is consistent with the menstrual cycle pattern, a repeating transition between SE cell states during the cycle. The median latent time was much shorter during the secretory phase, indicating the export of mature mRNA to the cytoplasm occurs faster during the secretory phase (**Fig. 5J** and **S4F**). SE cells were more transcriptionally active during the secretory phase, consistent with the *CellPhoneDB* analysis (**Fig. 5H**).

### Secretory epithelial cell markers can subtype high grade serous ovarian cancer

Given the mounting evidence suggesting FT epithelial cells are the cell of origin of HGSC (26), we investigated how various FT epithelial cell markers identified in our study are represented in various cancers of The Cancer Genome Atlas (TCGA) dataset that reported bulk RNA-seq data of various cancers (27). We focused on the top differentially expressed genes that we identified in SE cells from pre-menopausal (SE1/2/3-pre), and post-menopausal samples (SE-post, SE-post-specific) and performed a deconvolution analysis of the different TCGA molecular subtypes of ovarian, breast, head and neck, and lung squamous cell carcinoma (**Fig. 6A**).

**Figure 6:**
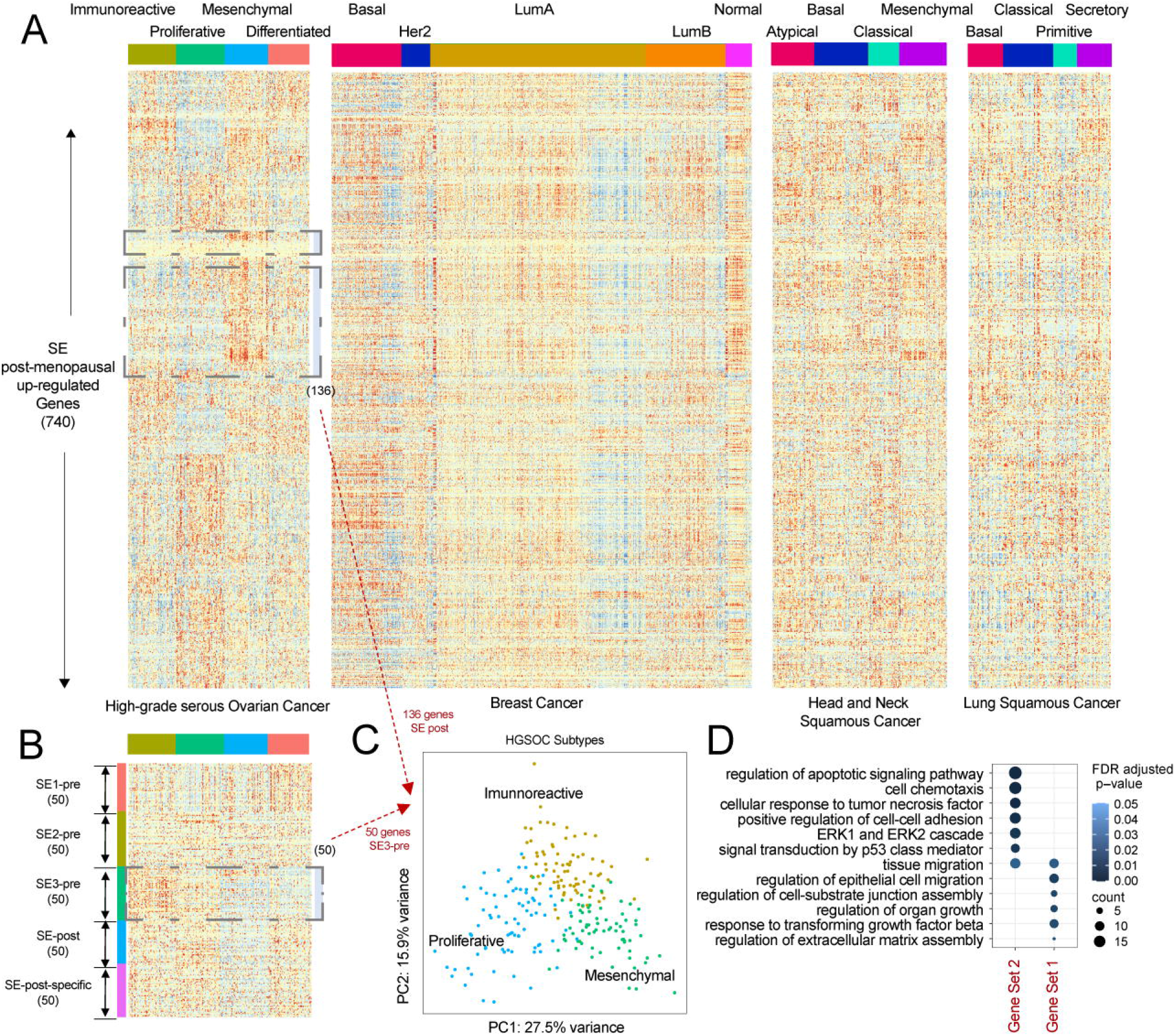
Characterization of molecular subtypes of ovarian cancers using secretory epithelial cell markers. A) Heatmaps showing the top 740 up-regulated genes in our post-menopausal SE cells across the various molecular subtypes of high-grade serous ovarian cancer, breast cancer (BRCA), head and neck squamous cell carcinoma (HNSC), and lung squamous cell carcinoma (LUSC) as described by TCGA. B) Heatmap showing the top 50 expressed markers in pre- and post-menopausal SE subtypes across the ovarian cancer molecular subtypes as identified by TCGA. C) Principal component analysis plot for TCGA samples (n= 304) using 186 SE markers (SE3; n = 50 genes, and SE post-menopausal; n = 136 genes). D) Pathway enrichment analysis of selected SE marker sets derived from post-menopausal SE (gene set 1) and SE3 (gene set 2).

The SE markers upregulated in post-menopausal women (740 genes, **Table S12**) adeptly recapitulated the previously described molecular subtypes of HGSC: immunoreactive, proliferative, mesenchymal and differentiated (27). A subset of 136 genes (**Table S12**) displayed high expression in HGSC of the mesenchymal subtype (**Fig. 6A**). Upon further scrutiny of the top expressed markers found in the different pre- and post-menopausal SE cell types, we discerned that the top 50 SE3-pre gene markers (**Table S12**) were prominently expressed in the immunoreactive TCGA HGSC subtype (**Fig. 6B**). By juxtaposing these patterns with those from other cell types and TCGA cancer types, we were able to confirm that these associations are uniquely tethered to the SE cells in ovarian cancer. We combined the top 50 SE3-pre gene markers associated with immunoreactive-(**Fig. 6B**), and the 136 SE-post markers associated with mesenchymal HGSC in the post-menopausal FT and performed principal component analysis. Strikingly, the 186 markers effectively separated immunoreactive, proliferative, and mesenchymal TCGA HGSC subtypes, but not the differentiated subtype (**Fig. 6C**, **S5A**). The 136 SE-post genes associated with the mesenchymal subtypes showed enrichment in cell migration and cell junction assembly (**Fig. 6A**). The 50 SE3-pre gene markers, associated with the immunoreactive subtype (**Fig. 6B**) exhibited enrichment in apoptosis, chemotaxis, response to tumor necrosis factor, and cell adhesion (**Fig. 6D**). The immunoreactive subtype as identified by the 50 SE3-pre genes demonstrated notably better survival compared to proliferative and mesenchymal subtypes combined (**Fig. S5B**).

## Discussion

Our study presents compelling evidence that menopause leads to substantial changes in the cellular composition and gene expression profiles of the FT. While uterine and ovarian atrophy during menopause has been extensively described (28), our findings indicate that the FT also undergoes previously undescribed atrophic changes. These changes are characterized by alterations in cellular composition and gene expression that are indicative of aging and stress responses.

Moreover, our study delineates the molecular and transcriptional alterations occurring within FT cells throughout the menstrual cycle, especially for SE and ST cells. During the proliferative phase (estrogen^high^, progesterone^low^) of the menstrual cycle, SE2 cells dominate the FT epithelium. These cells express SOX9^+^ LGR5^+^ in SE2 cells and activate pathways involved in response to fibroblast growth-factor stimulation, while stromal cells express genes involved in cellular energy, proliferation, and metabolic processes. Correspondingly, in the uterus, the proliferative phase is characterized by the proliferation of endometrial cells, which leads to the thickening of the uterine lining (22). This menstrual cycle phase in the endometrium is characterized by SOX9^+^ LGR5^+^ expressing stem/progenitor epithelial cells (29). Given that the uterine and FT epithelium share the mesoderm as a common embryological origin, we speculate that the SE2 cells in the FT are specialized, highly proliferative stem cells with a marked response to estrogen.

We showed that SE1 cells dominate the FT epithelium during the secretory phase (estrogen^low^, progesterone^high)^ of the menstrual cycle. Elevated progesterone during this phase plays a crucial role in fostering immune cell tolerance, through the differentiation and expansion of Treg cells (30), to ensure successful fertilization, transport and implantation. We saw an increase in T/NK cells during the secretory phase of the menstrual cycle and found that SE1 cells upregulate pathways involved in responses to steroid hormone regulation and MHC class II assembly and processing, indicating menstrual cycle-dependent regulation of immune cells (30, 31). Progesterone alters the stromal cell composition and influences epithelial-stroma crosstalk during embryo implantation in the uterus. In alignment with this role of progesterone, our findings indicate an enhancement in epithelial-stromal communication during the secretory phase (32). Collectively, these findings suggest that gene expression in various cell types within the uterus and FT is likely regulated in a similar manner throughout the menstrual cycle.

The FT epithelium is probably the site of origin for most HGSC (26, 33). The earliest event of malignant epithelium transformation is marked by the outgrowth of secretory cells and acquisition of aberrant TP53 protein, also known as the p53-signature (4). Herein, we identify a post-menopausal specific SE cell cluster (‘SE-post specific’), with increased TF activities of p53 family members and RUNX3. Notably, RUNX3’s role as a tumor suppressor or oncogene is contingent upon the p53 status, and it is implicated in DNA damage repair pathways (34). Indeed, we observed enrichment in p53-mediated signal transduction and DNA damage response. IHC confirmed the presence of nuclear p53 positive cells in the FT epithelium, suggesting that this could be indeed an early transformation event. However, it remains undetermined whether these cells possess malignant potential or are destined for apoptotic cell death. The TCGA HGSC data (27) categorized four molecular subtypes of HGSC of the ovary: proliferative, immunoreactive, differentiated and mesenchymal. We showed that SE-postmenopausal upregulated genes and SE3-premenopausal markers, can be used to successfully classify the proliferative, immunoreactive and mesenchymal subtypes of HGSC. Previous work by Hu et al., 2020 uncovered the differentiation trajectory of post-menopausal FT epithelial cells, revealing four FT secretory epithelial cell subtypes that are linked to HGSC cell states (13). This supports the theory that SE cells may be the progenitors of HGSC, particularly for the immunoreactive, proliferative, and mesenchymal subtypes, suggesting an alternative origin for the differentiated subtype. (35, 36).

In summary, we have systematically characterized gene and TF expression in pre- and postmenopausal women and during different menstrual cycle phases. These datasets will help to start exploring the biological functions of the FT and explain why certain diseases are more prevalent in pre-menopausal, while others are more common in post-menopausal women.

## Limitations of the study

We did not measure blood hormone levels to determine menstrual cycle phases but determined them based on patient history and endometrial pathology. We reported three patients in secretory phase, four patients in proliferative phase and 3 patients of unknown phase due to use of contraception. Therefore, analysis were performed on combined cells from all anatomical regions (A and F) for scRNA-and scATAC-seq samples. For similar reasons, and due to the limited number of pre-menopausal isthmus (I) samples collected, we did not include them into the menstrual cycle analysis.

scATAC-seq was only performed on a subset of donors used for scRNA seq (D14 – 18), resulting in fewer cell numbers sampled. As a result, we were not able to identify all cell types, identified in scRNA-seq, in our scATAC-seq analysis.

## Data Availability

De-identified human single-cell RNA and ATAC sequencing data have been deposited at CELLxGENE and at the European Genome-Phenome Archive (EGA). They are publicly available as of the date of publication. The link to the data is listed in the key resource table.

## Acknowledgements

We thank the women who donated the tissues and the surgeons that helped make this study possible. We also thank the Human Tissue Resource Center, the Genomics Facility, and the patients that kindly donated tissues for this study.

## Author contributions

E.L., A.B., and M.C. jointly oversaw project design and analysis. Computational analyses were performed by Y.L., L.Z., and M.C. Tissue dissociations, and 10X Genomics single-cell RNA and ATAC-seq assays were performed by H.E., P.B., S.A., R.K. and M.W, under A.B.’s guidance. M.W. performed Visium (by 10x Genomics) experiments, immunofluorescence, and immunohistochemistry. M.W. designed all images made with Biorender.com. P.B. and M.W. performed all tissue isolations and cultures for *in vitro* studies. R.L. and A.J.B. read histology and immunohistochemistry. S.A., P.B. and R.K. consented all women donating tissues. E.L., A.B., M.W., Y.L., and M.C. wrote the manuscript. All authors reviewed and agreed on the final version of the submitted manuscript.

## Methods

### Resource Availability

#### Lead contact

Further information and requests for resources should be directed to and will be fulfilled by the lead contact, Mengjie Chen.

#### Materials availability

Requests for resources and reagents should be directed to and will be fulfilled by the lead contact, Mengjie Chen.

#### Data and code availability

This paper does not report original code.

Any additional information required to reanalyze the data reported in this paper is available from the lead contacts upon request.

### Experimental Model and Subject Details

#### Human subjects and tissue acquisition

We collected fallopian tube fimbria (F) and ampulla (A) samples from 10 pre-menopausal women under the age of 50, who underwent surgeries for fibroids, prolapse or elective sterilization. A gynecologic pathologist (R.L., A.J.B.) determined the menstrual cycle phases for each patient. Phases were determined based on the microscopic appearance of the normal endometrium and abstracted from the pathology report (**Table S1**). We obtained informed consent from all study participants prior to their surgery. All procedures were conducted under the approval of the Institutional Review Board at the University of Chicago.

### Method Details

#### Immunohistochemistry

We used formalin-fixed and paraffin-embedded tissue blocks from samples collected as part of this study. Briefly, five µm sections were stained with hematoxylin and eosin or with commercially available antibodies using the Leica Bond RX automated stainer (Leica Biosystems) using the following antibodies: OVGP1 at 1:500 (Thermofisher Scientific, PA5-64007), pantropic p53 at 1:100.

#### Tissue dissociation, primary cell isolation and hormone treatments

We collected and individually processed all pre-menopausal ampulla and fimbria samples as previously described (11), with the exception that the digestion time for epithelial and stromal cells was reduced from 30 minutes to 15 minutes.

For *in vitro* studies, FTECs were isolated and cultured as previously described (37). Following epithelial cell removal, we used the remaining stromal and fibroblast fraction for further processing. The stroma and fibroblast fraction was rinsed in PBS and digested (12-18h) in collagenase type III (1000 U, Worthington Biochemical, LS004206) in 10% fetal bovine serum (FBS, Applied biological materials, TM997-100) in DMEM (Thermo Scientific, A1443101). Following digestion, cells were centrifuged, and the resulting pellet washed in PBS. Fallopian tube stromal cells (FTSCs) were plated in DMEM and 10% FBS.

The FTECs were plated at 250,000 cells on primaria coated 6-well tissue culture plates (Corning, 353846) and the FTSCs were plated at 200,000 cells on 6-well tissue culture plates. Both FTECs and FTSCs were cultured at 5% CO_2_, 37°C for 24-48 hrs or until 70% confluence was achieved. Cells were then primed with 500 nM β-estradiol (E2, Sigma-Aldrich, E2758), medroxyprogesterone 17-acetate (P4, Sigma-Aldrich, M1629) or DMSO (Control) for 8 hours before cell were harvested and RNA isolated.

#### Immunofluorescence

Chamber slides were coated with fibronectin (5 µg/mL, BD biosciences, 356008). Human FTEC and FTSC cells were plated at 70,000 and 30,000 cells per well respectively. Cells were tretaed with 500 nM medroxyprogesterone 17-acetate or 500 nM β-estradiol for 8 hrs, fixed with 4% paraformaldehyde, followed by permeabilization with 0.5% Triton X-100 and blocking with goat serum. Slides were stained overnight at 4 °C in goat serum using the following primary antibodies: IGF1 (1:200, OriGene Technologies, TA805748S) and IGF2 (1:200, ThermoFisher scientific, MA532485), followed by fluorescently labeled secondary antibodies (1:200, Alexa Fluor 488 and 568, Invitrogen) and Hoechst 33258 (1:200, Molecular probes, H-3569) for one hour. We imaged slides on a Nikon Eclipse Ti2 microscope and processed images with NIS-Elements (Nikon).

#### 10X Genomics 3’ Single-cell RNA sequencing

Samples from 10 pre-menopausal women were used for 10x Genomics 3’ scRNA-seq. Single-cell suspensions were prepared as described above, washed and re-suspended to achieve a target cell count of 700-1200 cells/µl. All 3’ scRNA-seq libraries were generated as described previously (11) targeting 8,000 cells per sample. We sequenced the resulting libraries using the Illumina NextSeq 500, (using 75 cycle kits) or Novaseq 6000 (using 100 cycle kits) at 30,000-50,000 reads per cell.

#### Single-cell RNA sequencing data analysis

We converted and demultiplexed raw BCL (binary base call) sequencing data into FASTQ files as described previously (11). In brief, raw sequencing reads were aligned to human genome reference hg38, filtered, and quantified as unique molecular identifier (UMI). We refined the UMI count matrix by applying a systematic analysis workflow (scRICA) with the following quality control (QC) criteria: 1) cells must express at least 200 gene features, and each gene feature must be present in a at least 3 cells; 2) removal of doublets and triplets as identified by *DoubletDecon*; and 3) filtering of low-quality cells with ≥20% mitochondrial content. Next, UMI count matrices were log-normalized and cells from different samples integrated. Unwanted variations were eliminated prior to dimensionality reduction using PCA and construction of a shared nearest-neighbor (SNN) graph.

We then compared the cell clusters obtained from unsupervised learning to our post-menopausal study, and manually annotated them using literature curated canonical cell type markers (**Table S3**). We used HIPPO to refine the subtypes of NK/T and SE. Premature termination was prevented by applying a z-score threshold of 1 and maintaining the default outlier proportion at 0.001% in HIPPO. The resulting NK/T and SE sub-clusters were further characterized using additional gene marker lists.

#### Ligand–receptor analysis with CellPhoneDB

We utilized normalized gene expression data from six pre-menopausal donors with clearly identified menstrual cycle phases as input for ligand-receptor analysis. The CellPhoneDB database v2.0.0 (https://www.cellphonedb.org/) was employed to detect ligand-receptor interactions (38). For each of the two distinct menstrual cycle phases, we determined significant ligand–receptor interactions between any pair of cell types, using a statistically significant level of 0.05. To examine and compare the differences between the menstrual cycle phases, we chose the ligand–receptor interactions that were most closely related to hormone response.

#### Functional enrichment analysis

For functional enrichment analysis, we used various identified gene sets as input files, including SE cells from different menstrual cycle phases, SE cells from pre- or post-menopausal patients, and the top 200 over-expressed genes in each sub-cluster of SE and ST cells. We analyzed these gene sets with the Gene Ontology (GOs) database, concentrating on three functional categories: biological processes (BP), cellular components (CC), and molecular functions (MF). The entire analysis was conducted in R using the bioconductor package *clusterProfiler*. Enriched functions were identified at FDR-corrected p-values of 0.1, and we presented selected GO terms in dot-plots for visualization.

#### Velocity analysis

We used velocyto (version 0.2.5) (https://velocyto.org/) to obtain spliced and unspliced count matrices for each cell in our scRNA-seq dataset (24). We used these matrices to estimate the rate of splicing and transcription for each gene in individual cells. We used BAM files of aligned reads for each sample as input for velocyto. Then, the unspliced and spliced matrices of each sample w read and merged into an AnnData object for single-cell data storage and manipulation. For downstream analyses we divided cells into subsets based on their assigned cell-and phenotype. The velocity stream and latent time were computed for each cell in the dataset using the dynamical model. The velocity stream represents the direction and magnitude of the change in RNA over time. The latent time represents the time elapsed since the start of the transcriptional program.

#### Single-cell ATAC sequencing

We performed scATAC-seq as previously described (11). Briefly, single-cell suspensions were lysed to obtain intact nuclei, followed by nuclei tagmentation to generate scATAC-seq libraries. For scATAC-seq we targeted 8,000 cells with 6,000-8,000 nuclei per sample. We used Agilent 2100 Bioanalyzer traces to evaluate library quality and performed sequencing with the University of Chicago Core Facility on the Illumina NextSeq 500 or NovaSeq 6000 platforms. Libraries were sequenced at 25,000-30,000 (at PE75 and PE100, respectively) per nucleus.

#### Pre-processing and quality control of scATAC-seq data

We performed scATAC-seq data analysis as previously described (Lengyel et al., 2022). In brief, raw sequencing data were aligned to human hg38 reference genome and R Seurat v4.0 and *Signac v1.4.0* (39) used for further analysis. Quality control metrics for samples to be included in term-frequency inverse-document-frequency (TFIDF) normalization were: 1) cells with peak region fragments between 1,000 and 20,000; 2) reads in peaks >15%; 3) blacklist ratio < 0.05; 4) nucleosome signal < 10; and 5) TSS enrichment > 2. Next, we performed label transfer (from scRNA-seq) and merged Seurat objects from each sample based on the common peak set created by merging peaks from all the datasets. Dimensional reduction was performed, and batch effects were corrected on the first 25 latent semantic indexing (LSI) components. The first LSI was excluded because it was highly correlated with sequencing depth. Gene activity scores were estimated and corrected data were used for unsupervised clustering analysis.

#### Integration of scRNA-seq and scATAC-seq datasets

We used cells from matched scRNA-seq as a reference to predict and obtain cell types in our scATAC-seq data as previously described (Lengyel et al., 2022). In short, the variable features of the scRNA-seq data were used as reference, while the scATAC-seq data was used as the query data to generate the gene activity matrix. Next, transfer anchors were learned, and cell type labels predicted. We assigned each cell in the scATAC-seq with a cell type identity from the matching scRNA-seq data. We only retained cells with a prediction score > 0.5 for further analysis. Further cell type sub-clusters were transferred, if supported by the unsupervised scATAC-seq data. We performed label transfer for each patient separately.

#### Transcription factor motif analysis

Transcription factor (TF) activities were estimated as previously described (Lengyel et al., 2022). Briefly, TFs and their binding motifs listed in cisBP (14) were used as another input to chromVAR (40) for positional weight matrix calculation. We applied RunChromVAR in Signac to calculate cell type-specific TF activities. Differential activities for each cell type by menopausal status were computed with *FindMarkers* using Bonferroni-adjusted p-values < 0.05. We performed TF enrichment analysis of all motifs for each cell separately. We then compared and displayed the results.

#### Tissue preparation and image acquisition for spatial transcriptomics

Fresh tissues were embedded in cryomolds with ice-cold Tissue Tek O.C.T (FisherScientific, 4585) and the blocks stored at −80°. Prior to the placement of 10µm frozen sections onto the gene expression and tissue optimization slides (10x Genomics), we analyzed RNA Integrity Number values to determine RNA quality. Tissues were optimized according to the manufacturers user guide (Visium spatial gene expression reagents kits-tissue optimization; document number: CG000238 Rev D). The slides and samples were processed as described by the manufacturer user guide for frozen samples (Visium spatial gene expression reagent kit, document number: CG000239 Rev E) and libraries generated. Samples were sequenced on the Illumina NovaSeq using 75 bp PE.

#### Quantification and Statistical Analysis of TCGA data

We summarized cell numbers based on cell annotations and normalized the relative cell percentage by the total number of cells. We only reported significance with p < 0.05.R *survival* and *survminer* packages was used for survival analysis on the TCGA OV cohort. We used the Kaplan Meier method to generate survival curves and used log rank test to compare the survival curves regarding gene expression levels.

Unless stated otherwise, quantitative data are represented as mean ± standard deviation. We indicated sample sizes, statistical tests, and p values in the figure legends. Statistical analysis of qPCR results was performed with GraphPad Prism 8 software. One-way ANOVA comparison of the mean was used to determine significance.

## Tables

**Table S1: Patient cohort and clinical-pathologic information.**

**Table S2 (related to Figure 2): Cellular composition of pre- (isthmus, ampulla, and fimbria) and pre-/post-menopausal (ampulla and fimbria) fallopian tubes (scRNA-seq).**

First tab: Cellular composition for each cell cluster for pre- and post-menopausal ampulla and fimbria combined.

Second tab: Cellular composition for each cell cluster for pre-menopausal fallopian tubes by anatomical site (ampulla, isthmus, and fimbria).

Third tab: Cellular composition for each cell cluster for post-menopausal fallopian tubes by anatomical site (ampulla, isthmus, and fimbria).

**Table S3 (related to Figure 2): Canonical marker genes used to annotate cell types in the fallopian tube (scRNA-seq).**

First tab: canonical cell marker genes.

Second tab: Marker genes for fallopian tube epithelial and PEG cells.

**Table S4 (related to Figure 2): Total cell numbers and relative percentages of individual cell clusters found in the pre-menopausal fallopian tube by patient (scRNA-seq).**

**Table S5 (related to Figure 2): Label transfer statistics between scRNA-seq and scATAC-seq standalone clusters.**

First tab: Label transfer summary for each sample.

Second tab: Cell type composition by cell type for pre- and post-menopausal samples.

**Table S6 (related to Figure 3): Transcription factor activities in pre-menopausal fallopian tubes by cell type.**

**Table S7 (related to Figure 4): Differential gene expression results for pre- and post-menopausal SE cells.**

**Table S8 (related to Figure 4):): Differential transcription factor activities in pre- and post-menopausal fallopian tubes by cell type.**

**Table S9 (related to Figure 4): Top 100 overexpressed genes and selected, associated gene ontology for all SE sub-types.**

First tab: Top 100 overexpressed genes for each secretory epithelial subtype.

Second tab: Selected gene ontology enrichment terms for each secretory epithelial subtype.

**Table S10 (related to Figure 5): Gene ontology enrichment analysis of pre- and post-menopausal stromal cells.**

**Table S11 (related to Figure 5): Differentially expressed genes in SE cells during the secretory and proliferative phase using pseudo-bulk analysis.**

**Table S12 (related to Figure 6): Gene markers upregulated in post-and pre-menopausal SE cells.**

First tab: 740 SE marker genes upregulated in post-menopausal donors.

Second tab: 136 SE marker gene subset with high expression in mesenchymal HGSC. Third tab: Top 50 SE3-pre marker genes.

## Supplementary Figure Legends

**Figure S1** (related to Figure 2): **Cellular composition of the normal pre-menopausal fallopian tube.**

A) Primary cell types found in both the isthmus (n = 5), ampulla (n = 10), and fimbria (n = 10) of the normal pre-menopausal fallopian tube. The UMAP plot visualizes the 12 major cell clusters identified using scRNA-seq. Cell types are abbreviated as follows: SE = secretory epithelial cells; CE = ciliated epithelial cells; T/NK = T and natural killer cells; MP = macrophages; MA = mast cells; ST = stromal cells; SM = smooth muscle cells; LE = lymphatic endothelial cells; EN = endothelial cells; P/V = pericytes and vascular smooth muscle cells; B/P = B cells and plasma B cells.

B) Relative abundance of the 12 major cell clusters identified in the pre-menopausal fallopian tube by anatomic site using scRNA-seq. The graph shows the individual percentage of each cell type for all donors combined.

C) UMAP plot profiling of (A) identifying 11 major cell clusters in the fallopian tube by anatomical site (ampulla: n = 5, isthmus: n = 3, and fimbria: n = 5). The labels are transferred from scRNA-seq data and scATAC-seq specific cluster labels are added.

D) Heatmaps based on scATAC-seq data. Transcription factor activity by cell type in the different fallopian tube anatomical region. Heatmap includes all 869 motifs available in the cisBP database.

**Figure S2** (related to Figure 3): **Menopause changes the cellular composition and gene expression profiles of cell types found in the normal human fallopian tube.**

A) Dot plot showing combined (ampulla and fimbriae; pre-menopausal donors: n = 10; postmenopausal donors: n = 7) normalized gene expression levels of known canonical marker genes for each cell type identified in the pre- (n = 20 sample sites) and post-(n = 12 sample sites) menopausal fallopian tube excluding SE cells.

B) Volcano plot derived from pseudo bulk analysis for combined anatomical sites (ampulla and fimbria). The volcano plot is showing the differential gene expression analysis of genes expressed in ST1-3 based on Menopausal status.

C) Heatmaps showing the frequency of interactions among different cell types in the pre- and post-menopausal ampulla (n = 6), and fimbriae (n = 6) imputed by CellPhoneDB from scRNA-seq.

**Figure S3** (related to Figure 4): **Menopause induces significant changes in gene regulation and -expression in epithelial cells.**

A) UMAP plot profiling of combined (ampulla and fimbria) fallopian tube samples by menopause status based on scRNA-seq data. UMAP plot identified 19 cell clusters in the pre-menopausal fallopian tube (n = 20 patient samples), while 18 cell clusters were identified in the post-menopausal fallopian tube (n = 12 patient samples). Boxes highlight changes in SE cell-clusters specific to menopausal status. SE 1/2/3 pre are specific to pre-menopausal fallopian tubes, while “SE post-specific” cells are specific to postmenopausal women.

B) Dot plot showing combined (ampulla and fimbriae) normalized gene expression levels of known canonical marker genes for SE sub-types identified in the pre- and post-menopausal fallopian tubes.

C) Dot plot showing normalized hormone receptor gene expression levels in SE subtypes by anatomical site. A = ampulla, F = fimbria.

D) Pantropic p53 immunohistochemistry in patient-matched fallopian tube epithelium. P53 staining is observed in the nuclei

E) Heatmap showing the average expression of TP53 consensus genes for SE cells at the sample level. Boxes highlight differences in consensus genes specific to SE sub-cell clusters.

**Figure S4** (related to Figure 5): **Epithelial cells undergo significant cellular and molecular changes during the proliferative and secretory phases of the menstrual cycle.**

A) Graphical figure of the menstrual cycle and hormone levels.

B) Cellular proportion differences (in percentages) of SE1 and SE2 cells based on anatomic region and menstrual cycle phase or progesterone-based contraception in pre-menopausal women. Proliferative phase (n = 4/4), secretory phase (n = 3/3), progesterone-based contraception (n= 3/3) for ampulla and fimbria respectively.

C) OVGP1 immunohistochemistry in the fallopian tube of patients on progesterone-based contraception (n = 2) and in menopause (n = 2).

D) Dot plot showing the enrichment of selected gene ontology (GO) cellular component terms. Terms are based on differentially expressed genes identified during the proliferative and secretory phases of the menstrual cycle using scRNA-seq data (FDR-corrected p-value <0.05).

E) Violin plots showing IGF1 and IGF2 gene expression in ST1 and ST2 cells during the proliferative-and secretory phase.

F) Latent time for SE 1-pre and SE2-pre cells.

G) Heatmap showing the number of interactions detected by *CellPhoneDB* (ampulla and fimbria combined) among different cell types in the fallopian tube by menstrual cycle phase.

**Figure S5** (related to Figure 6): **Characterization of molecular subtypes of ovarian cancers using secretory epithelial cell markers.**

A) Principal component analysis plot for TCGA samples (n = 304) for all four HGSC molecular subtypes (proliferative, mesenchymal, differentiated and immunoreactive) using our 186 identified SE-cell markers (SE3; n = 50 genes, and SE post-menopausal; n = 136 genes).

B) Survival plot for the immunoreactive molecular subtype against proliferative and mesenchymal subtypes combined.

## Abbreviations

A: ampulla
B/P: B and plasma B cells
CE: ciliated epithelial cells
EMT: epithelial to mesenchymal transition
EN: endothelial cells
F: fimbriae
FT: fallopian tube
FTEC: fallopian tube epithelial cells
FTSC: fallopian tube stromal cells
GWAS: genome-wide association studies
H&E: hematoxylin and eosin
HGSC: high-grade serous cancer
I: isthmus
IM: immune cells
LE: lymphatic endothelial cells
MA: mast cells
MN: monocytes
MP: macrophages
P/V: pericytes and vascular smooth muscle cells
scRNA-seq: single-cell RNA sequencing
scATAC-seq: single-cell assay on transposase-accessible chromatin
SE: secretory epithelial cells
SM: smooth muscle cells
ST: stromal cells
TCGA: The Cancer Genome Atlas
T/NK: T and NK cells
TF: transcription factors
SASP: senescence-associated secretory phenotype

