## Supplementary figures and images for "A Cellular atlas of the human fallopian tube reveals the metamorphosis of secretory epithelial cells during the menstrual cycle and menopause"

### Supplementary Figure 2

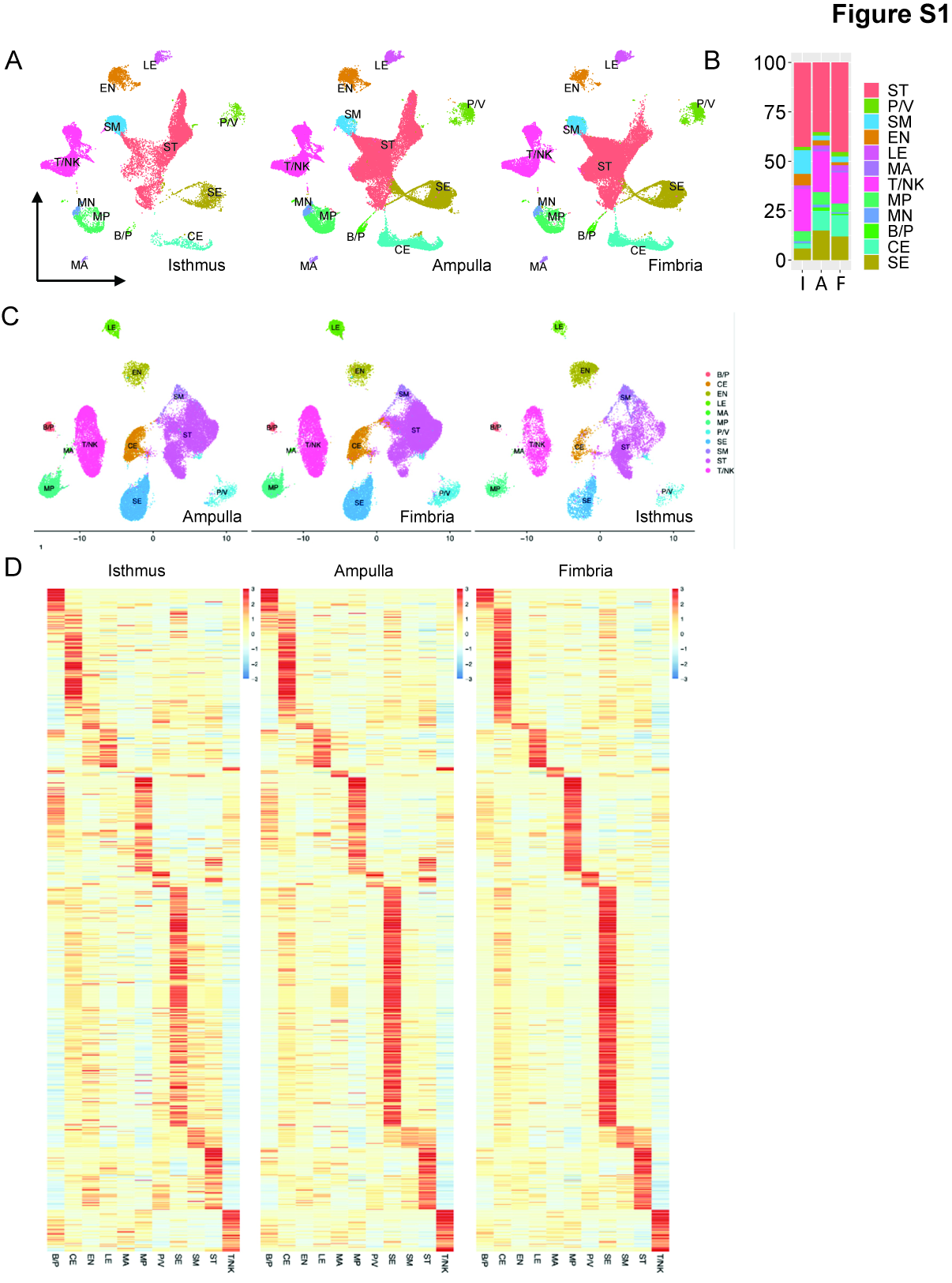

### Supplementary Figure 3

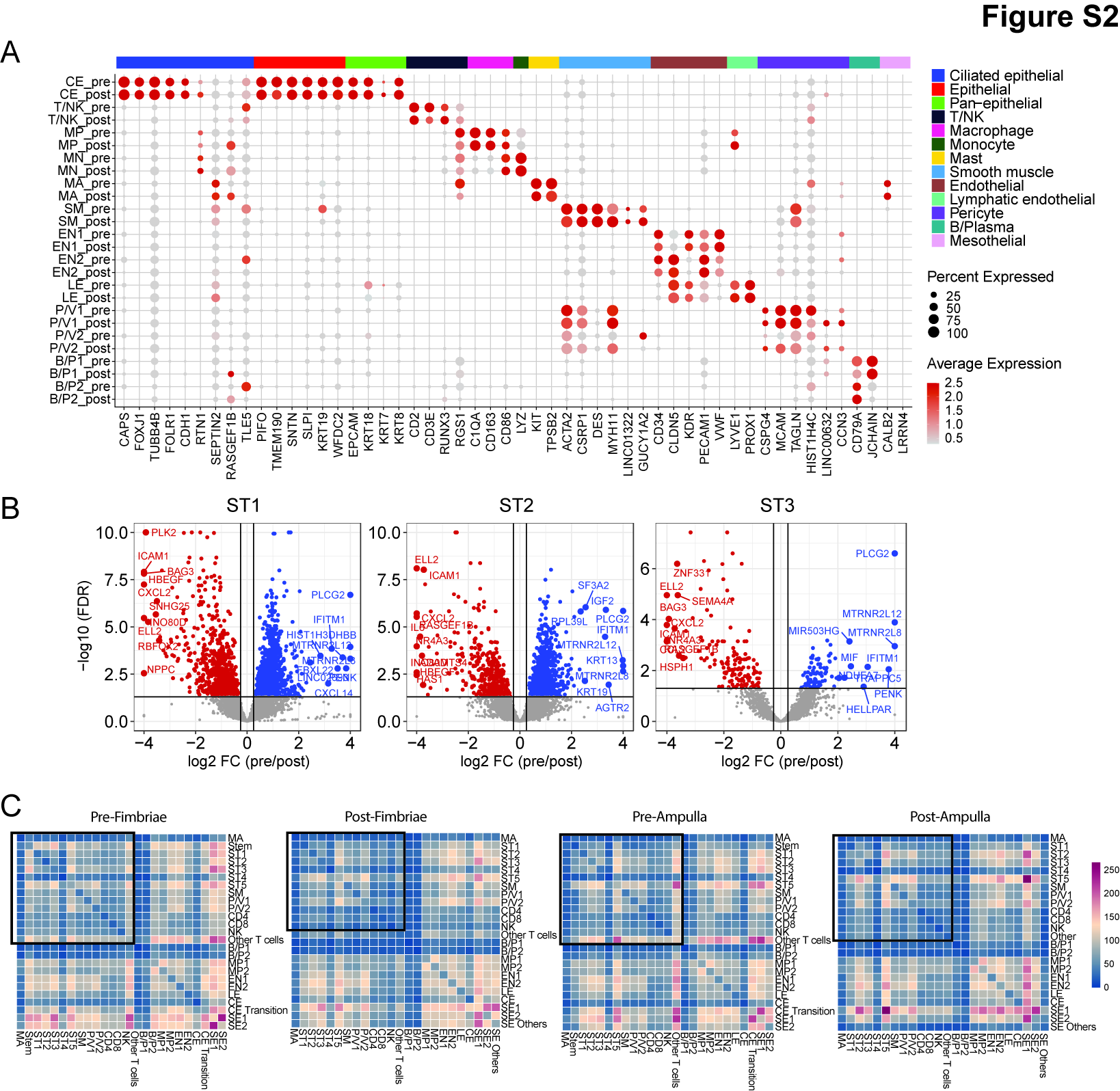

### Supplementary Figure 3

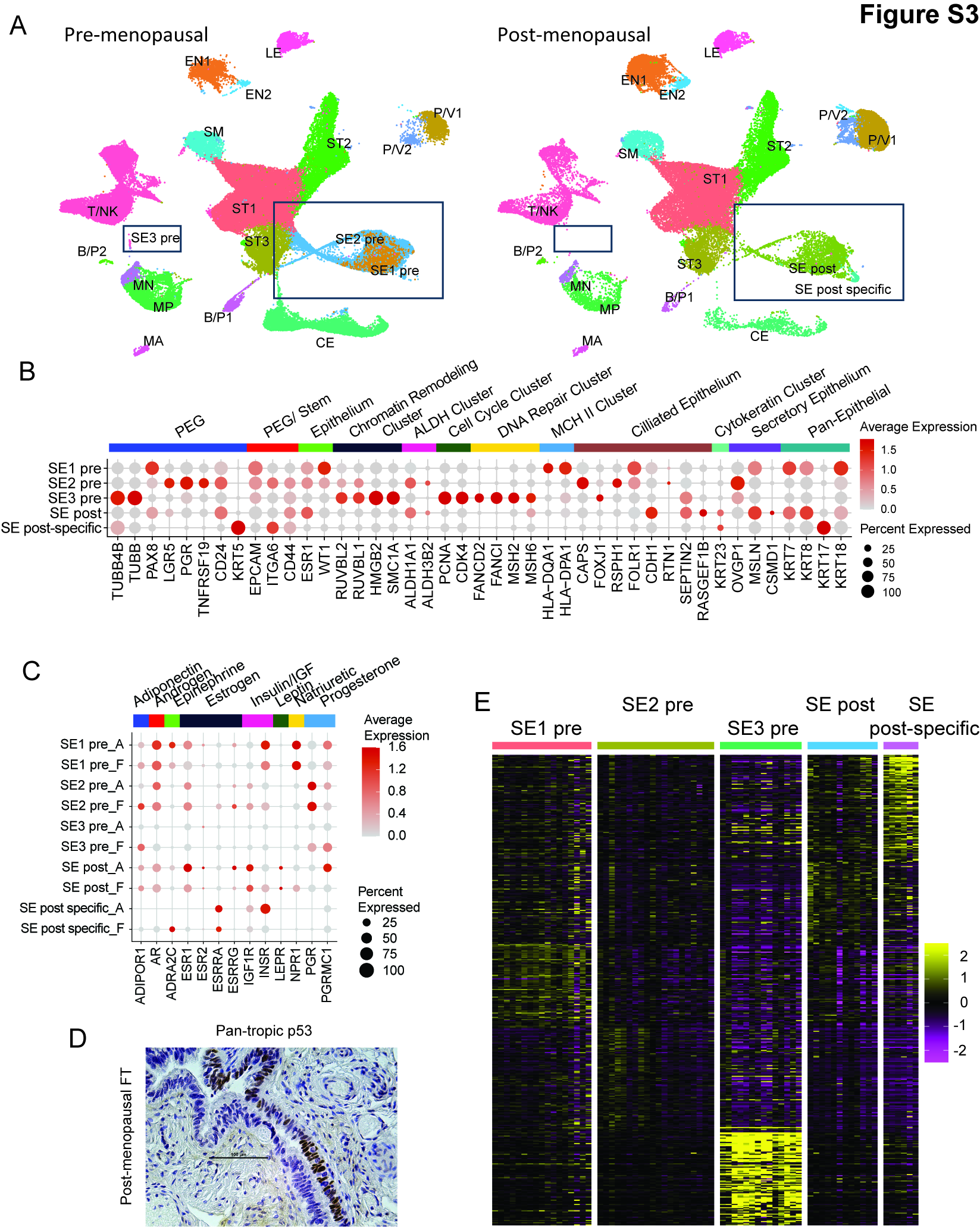

### Supplementary Figure 4

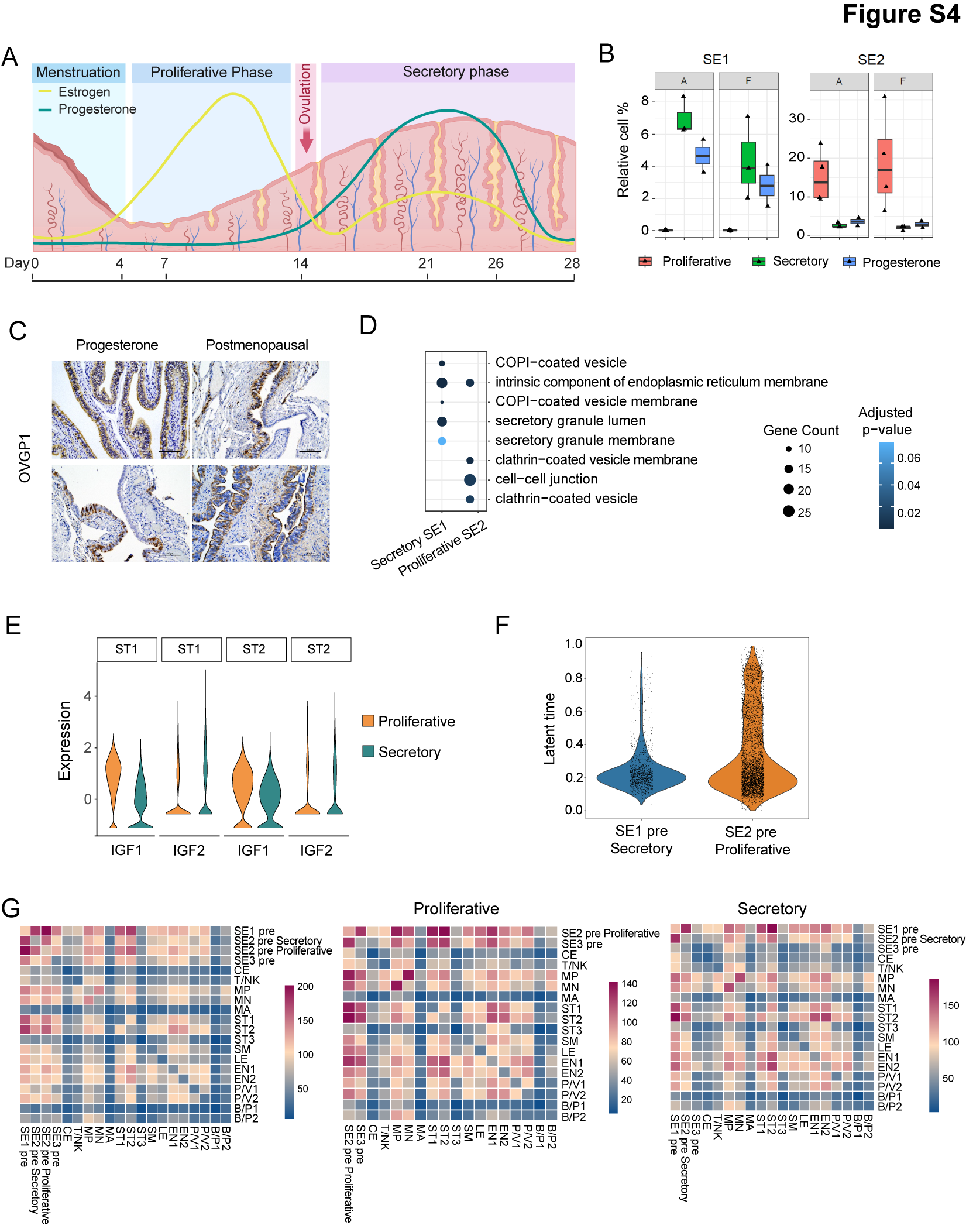

### Supplementary Figure 5

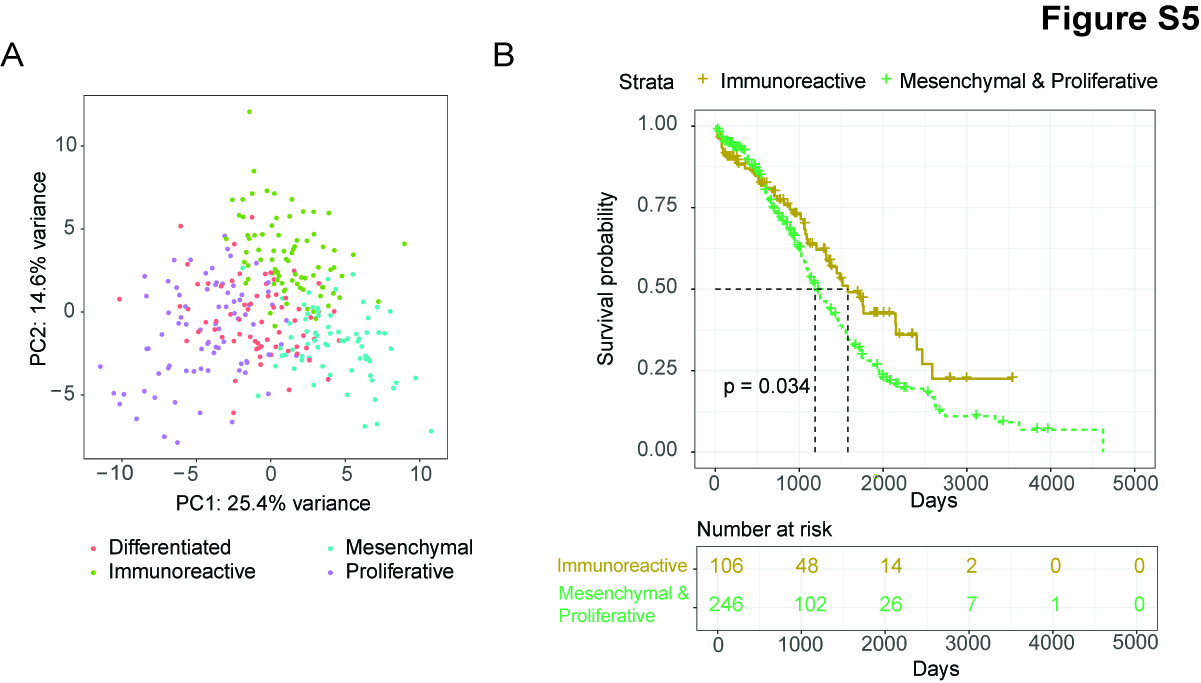
