## Supplementary Table 1 for "A Cellular atlas of the human fallopian tube reveals the metamorphosis of secretory epithelial cells during the menstrual cycle and menopause"

**Table S1 (related to Figure 1): Patient cohort.**

| **Sample ID** | **Age range (years)** | **Race** | **Hispanic** | **MP age**  **(years)** | **BMI** | **RNA-seq** | **ATAC-seq** | **Menstrual cycle** |
| --- | --- | --- | --- | --- | --- | --- | --- | --- |
| D1 | 66-70 | White | No | 51-55 | 27.58 | ^a^A, F, I, | - | Postmenopausal |
| D2 | 71-75 | White | No | 51-55 | 28.47 | ^a^A, I, | - | Postmenopausal |
| D3 | 61-65 | White | No | 56-60 | 31.48 | A, F, I, | A, F, I, | Postmenopausal |
| D4 | 51-55 | White | No | 51-55 | 21.45 | F, I | F, I | Postmenopausal |
| D5 | 61-65 | Asian | Unknown | 46-50 | 22.73 | A, F, I, | A, F, I, | Postmenopausal |
| D6 | 61-65 | White | Yes | 51-55 | 26.74 | A, F, I | A, F, I | Postmenopausal |
| D7 | 61-65 | White | No | 51-55 | 31.31 | A, F, | - | Postmenopausal |
| D9 | 46-50 | African American | Unknown | N/A | 26.67 | A, F | - | Secretory |
| D10 | 41-45 | African American | No | N/A | 53.34 | A, F | - | Proliferative |
| D11 | 36-40 | African American | No | N/A | 29.27 | A, F | - | Unknown/ Inactive endometrium^b^ |
| D12 | 41-45 | White | No | N/A | 28.03 | A, F, I | - | Unknown/ Inactive endometrium^c^ |
| D13 | 36-40 | White | No | N/A | 26.41 | A, F, I | - | Proliferative |
| D14 | 31-35 | African American | No | N/A | 32.33 | A, F | A, F | Proliferative |
| D15 | 41-45 | African American | No | N/A | 19.55 | A, F | A, F | Proliferative |
| D16 | 46-50 | African American | No | N/A | 33.11 | A, F, I | A, F, I | Unknown/ Inactive endometrium^b^ |
| D17 | 41-45 | African American | No | N/A | 41.01 | A, F, I | A, F, I | Secretory |
| D18 | 36-40 | African American | No | N/A | 28.62 | A, F, I | A, F, I | Secretory |

^a^Drop-seq data, ^b^ Depo-Provera, ^c^ Intra uterine device

All patients were non-smokers.

Abbreviations: donor (D), ampulla (A), isthmus (I), fimbria (F), ovary (O), Menopausal age (MP).
